# Beyond Usual Pace: Quick Walking Speed and Brain Health Outcomes in the Framingham Heart Study

**DOI:** 10.64898/2026.10.13.26365066

**Authors:** Anika Thomson, Sophia Lu, Vanessa M. Young, Haritha Vardhini Katragadda, Francis Kolo, Gabrielle Hromas, Monica Gireud-Goss, Mini E. Jacob, Joanne M. Murabito, Sudha Seshadri, Alexa Beiser

**Affiliations:** Glenn Biggs Institute for Alzheimer’s and Neurodegenerative Diseases, University of Texas at San Antonio Health Campus, San Antonio, TX; Department of Biostatistics, Boston University, Boston, MA; School of Social and Behavioral Sciences, Arizona State University, Tempe, Arizona; Swedish Health Services, Seattle, Washington; The Framingham Heart Study, Framingham, MA, USA; Department of Medicine, Boston University Chobanian & Avedisian School of Medicine, Section of General Internal Medicine, Boston Medical Center, Boston, MA, USA; Department of Neurology, Boston University Chobanian & Avedisian School of Medicine, Boston, MA, USA

**Keywords:** gait speed, dementia, stroke, aging, sex differences, executive function

## Abstract

**INTRODUCTION:** Stroke, dementia, and late-life depression share overlapping vascular pathophysiology, yet simple markers of shared neurological vulnerability across these conditions remain limited. Most gait research has relied on usual walking speed (UWS) alone, leaving it unclear whether quick walking speed (QWS) and gait adaptability (QWS minus UWS), relate to these conditions in the same way.

**METHODS:** We examined associations of UWS, QWS, and gait adaptability with executive function, verbal memory, depressive symptoms, and incident stroke and dementia in the Framingham Heart Study Offspring cohort. A larger sample (n=2,308; mean age 61.5±9.2 years; 54% women) was used for cross-sectional and stroke analyses, and a dementia-free sample of 1,148 participants aged ≥60 years (mean age 68.5±5.8 years; 53% women) was used for analyses of incident dementia. We estimated associations per SD slower walking or smaller adaptability using linear, logistic, and Cox regression (10-year follow-up), overall and in women and men.

**RESULTS:** Slower QWS and UWS were each associated with worse executive function, higher odds of high depressive symptoms (QWS OR 1.24, 95% CI 1.07-1.45; UWS OR 1.26, 95% CI 1.09-1.46), and greater hazard of incident stroke (QWS HR 1.30, 95% CI 1.03-1.64; UWS HR 1.36, 95% CI 1.11-1.65). Executive function and stroke associations were observed primarily in men (stroke QWS HR 1.55, 95% CI 1.11-2.17). For incident dementia, slower QWS reached significance (HR 1.34, 95% CI 1.10-1.63), while UWS did not (HR 1.20, 95% CI 0.98-1.46). Lower gait adaptability was associated with worse executive function overall and in women and men, and with incident dementia in women (HR 1.35, 95% CI 1.04-1.76), but not with depressive symptoms or stroke. No gait measure was associated with delayed recall.

**CONCLUSION:** Slower QWS was associated with incident dementia; UWS was not. Adding a brief quick walking trial to routine gait assessment may identify risk that habitual-pace screening overlooks.

**Key Points:**

- Quick-pace walking, which adds one instruction to the usual-pace test, was associated with higher 10-year dementia risk, whereas the usual-pace estimate was smaller and not statistically significant.
- The margin between quick and usual pace, which reflects the capacity to accelerate on demand, was associated with executive function and, in women, with incident dementia, but not with stroke or depressive symptoms, unlike either pace on its own.

**Why does this paper matter?:**

- Gait speed is among the most widely collected measures in geriatric practice and aging research, yet it is almost always recorded at usual pace alone. This study shows that the margin between quick and usual pace carries a different pattern of brain health associations than either pace on its own, so adding one timed trial to an assessment clinicians already perform makes available information that habitual-pace testing cannot provide.

## 1 INTRODUCTION

Dementia, stroke, and late-life depression collectively account for a substantial and growing share of age-related disability worldwide, contributing to more than 200 million disability-adjusted life years globally (1). These conditions arise in part from shared vascular and cerebral small vessel pathology and cluster within individuals, and estimates attribute at least 60% of strokes, 40% of dementia cases, and 35% of late-life depression cases to modifiable risk factors (1, 2). Identifying simple, low-burden markers that index neurological vulnerability before these conditions become clinically apparent could meaningfully advance early risk stratification in clinical and community settings.

Gait speed is a well-established marker of neurological and cardiovascular health in older adults (3-5). Most prior research has relied on usual walking speed (UWS) alone, which captures habitual locomotion, and community-based cohorts have linked slower UWS to cognitive decline and incident dementia (6, 7), stroke (8) and depressive symptoms (9). However, quick walking speed (QWS), which requires volitional acceleration, executive control, and cognitive-motor integration, may therefore expose neuromotor vulnerability that habitual pace masks. The difference between QWS and UWS, which we term gait adaptability and which others describe as walking speed reserve, indexes this capacity to modulate locomotion on demand (10). Prior work has examined this measure largely in relation to physical function and falls (11). In this cohort, slower QWS has been linked to incident clinically diagnosed Alzheimer’s disease dementia (12), although the two paces were not compared in the same participants. In the UK Biobank, maximal walking speed from wrist-worn sensors showed a stronger association with dementia than UWS (13). Few studies, however, have recorded both paces in the same participants and compared them, with their difference, across multiple brain-health outcomes.

Sex differences are well established across dementia, stroke, and depression, spanning disease burden, vascular risk, and brain structure (14, 15). Men accumulate cardiovascular and cerebrovascular burden earlier in life (16), while women show distinct patterns of brain connectivity and cognitive reserve (17). If quick walking speed and gait adaptability reflect the capacity to draw on reserve, these differences raise the possibility that the measures may carry different information in women and men (18). Yet few studies have examined gait-brain associations by sex, particularly across multiple outcomes.

To address these gaps, we examined associations of UWS, QWS and gait adaptability with cognitive performance (executive function and verbal memory), depressive symptoms, incident stroke, and incident dementia in the Framingham Heart Study (FHS), in the overall sample and separately in women and men. Because gait draws on executive and attentional control (4), we examined executive function and verbal memory separately. We hypothesized that (a) QWS would be more strongly associated with incident dementia than UWS, and (b) gait adaptability would show a similar pattern, consistent with quick walking capturing volitional gait capacity rather than absolute speed.

## 2 METHODS

### 2.1 Sample

We used data from the FHS Offspring (Gen 2) cohort, an ongoing community-based prospective study established in 1971 that enrolled 5,124 adult children of the Original Framingham cohort and their spouses (19). This study included Offspring participants who attended both (a) their seventh clinic examination between 1999 and 2005, and (b) a call-back examination (median 0.57 years later [IQR 0.29, 0.96]) at which walking speed testing and a neuropsychological battery were administered on the same day. We derived two analytic samples and defined baseline as the date of the call-back examination. The larger sample (n=2,308) included participants with complete gait, neuropsychological, depressive symptoms, and stroke data who were free of prevalent stroke and dementia at baseline and was used for cross-sectional analyses of neuropsychological performance and depressive symptoms, and for analyses of incident stroke. A second sample (n=1,148) was restricted to dementia-free participants aged ≥60 years at baseline who had *apolipoprotein* ε4 (*APOE* ε4) data and was used for analyses of incident dementia. Figures 1-2 present the sample selection process. This study was approved by the Institutional Review Board of Boston University Medical Campus/Boston Medical Center. All participants provided written informed consent.

**Figure 1.**
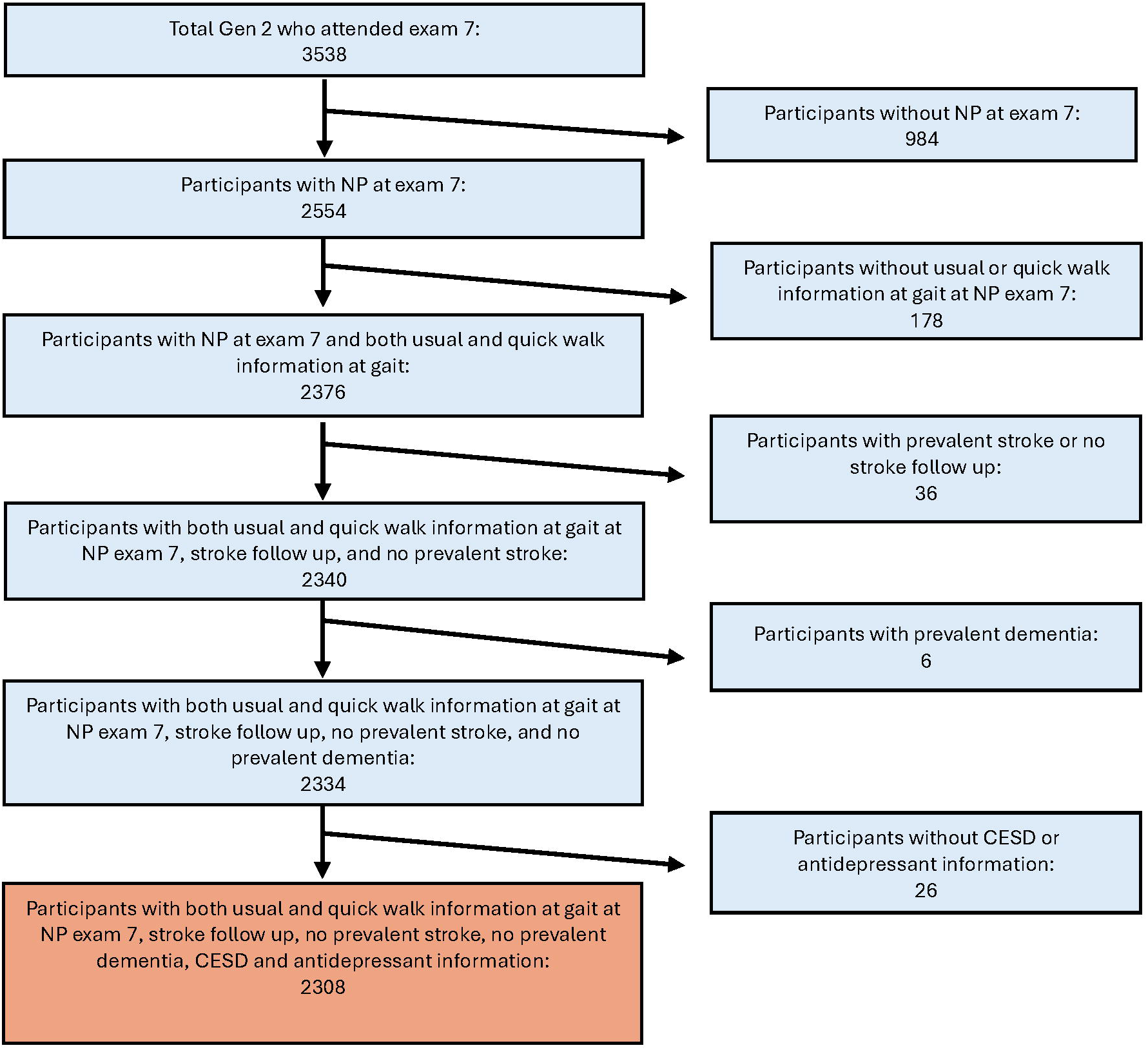
Sample Selection for the Stroke, Depression, and Neuropsychological Analytic Sample. Of 3,538 Framingham Heart Study Offspring (Gen 2) participants who attended the seventh examination cycle, 984 were excluded for missing neuropsychological data and 178 for missing usual or quick walking speed data. Of the remaining 2,376 participants, 36 were excluded for prevalent stroke or unavailable stroke follow-up data and 6 for prevalent dementia. A further 26 were excluded for missing depression data, yielding a final sample of 2,308 participants with complete gait, neuropsychological, stroke follow-up, and depression data. NP, neuropsychological assessment; CES-D, Center for Epidemiologic Studies Depression Scale.

**Figure 2.**
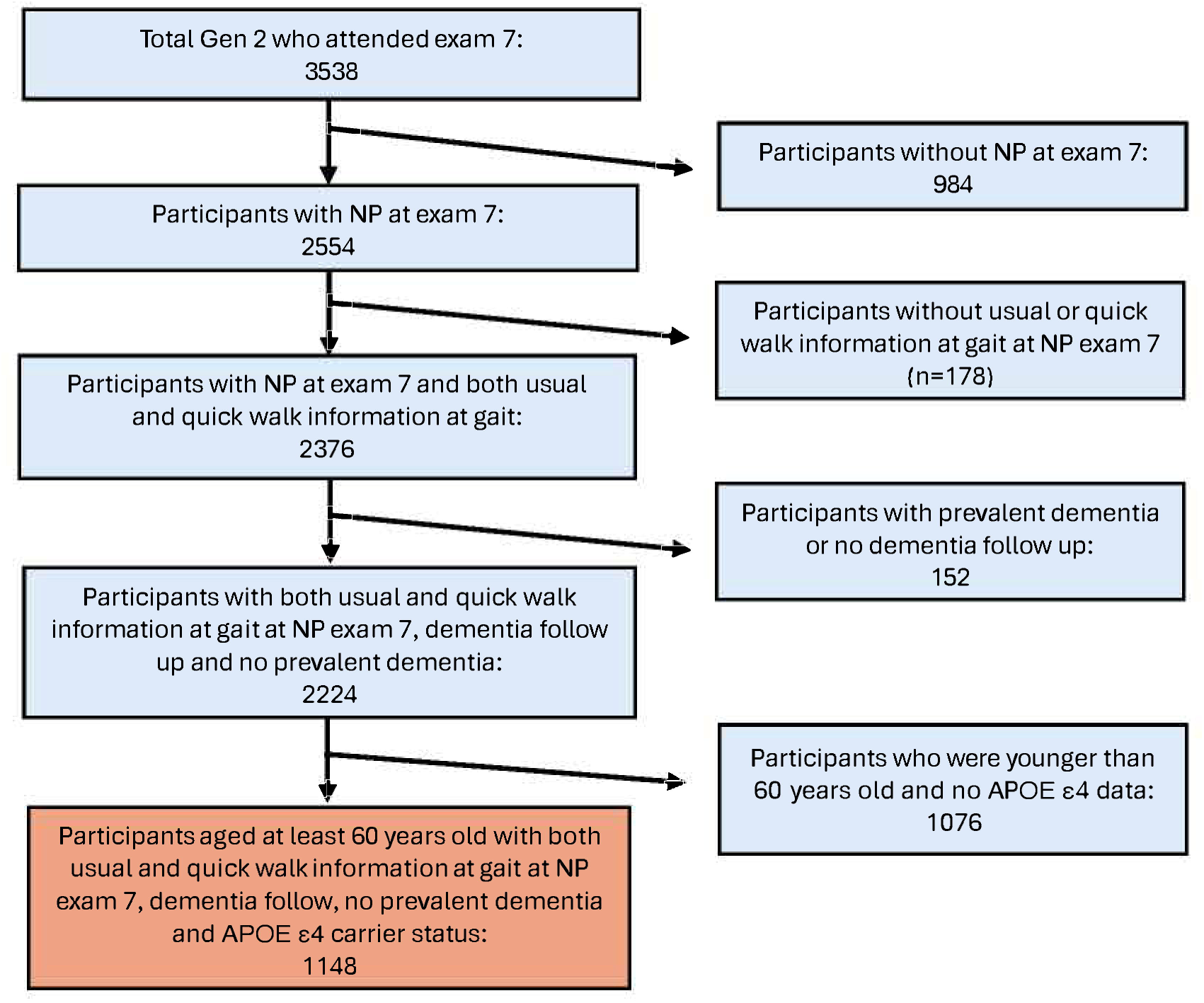
Sample Selection for the Dementia Analytic Sample. Of 3,538 Framingham Heart Study Offspring (Gen 2) participants who attended the seventh examination cycle, 984 were excluded for missing neuropsychological data and 178 for missing usual or quick walking speed data. Of the remaining 2,376 participants, 152 were excluded for prevalent dementia or unavailable dementia follow-up data. Of the remaining 2,224 participants, 1,076 younger than 60 years and without APOE data were excluded, yielding a final dementia analytic sample of 1,148 participants aged 60 years or older with complete gait, neuropsychological, and dementia follow-up data.

### 2.2 Gait Assessment

Trained research personnel assessed walking speed with a stopwatch as part of their motor testing battery. Participants walked a 4-meter course twice at their usual, self-selected pace, then covered the same course once as fast as possible without jogging or running (12). The time required to complete each course was recorded to the nearest second, and we converted these to speed in meters per second (m/s). We defined UWS as the faster of the two usual-pace trials and drew QWS from the single quick-pace trial. We calculated gait adaptability as QWS minus UWS. We log-transformed UWS and QWS to reduce skew, and added 0.341 to gait adaptability, which ranged from −0.33 to 2.67 m/s, before log-transforming it. We standardized each exposure and multiplied by −1 so higher values reflect slower walking or a smaller quick-to-usual difference.

### 2.3 Neuropsychological Assessment

Certified neuropsychological raters evaluated cognitive function using a standardized battery (20). We included the Trail Making Test Parts A and B (21), which assesses attention, processing speed, and executive function; and the Logical Memory Delayed Recall subtest of the Wechsler Memory Scale-Revised (22), which evaluates verbal learning and memory. We quantified executive function as the difference between completion times for Parts B and A (Trails B-A). We log-transformed Trails B-A (natural log) to reduce positive skew and multiplied it by -1 so that higher values on both tests reflect better performance.

### 2.4 Depression Symptoms

Participants completed the 20-item Center for Epidemiologic Studies Depression Scale (CES-D) during their seventh examination (23). We used an established cutoff score of ≥ 16 to indicate high depressive symptoms (24, 25).

### 2.5 Stroke and Dementia

#### 2.5.1 Stroke

The stroke surveillance protocol and procedures for determining stroke diagnosis and subtype have been described previously (26-28). A Framingham neurologist reviewed all potential stroke events, identified through hospital records, death certificates, and biennial participant contact, with final diagnosis informed by neuroimaging, clinical history, and examination findings. We defined stroke as an acute-onset focal neurological deficit of vascular origin persisting more than 24 hours, consistent with World Health Organization criteria, and included both ischemic and hemorrhagic subtypes (26-28).

#### 2.5.2 Dementia Surveillance and Diagnosis

Cognitive status was monitored at each clinical examination using the Mini-Mental State Examination (MMSE) (29-31). Participants were flagged for further evaluation through multi-source surveillance approach that integrated education-adjusted MMSE performance, prior FHS examination data, self- and family-reported cognitive concerns, physician referrals, medical records, and neuropsychological testing (29-31).

A multidisciplinary panel reviewed suspected dementia and established diagnosis, subtype, and estimated diagnosis date using serial clinical evaluations, caregiver interviews, medical records, neuroimaging, and autopsy findings where available. Dementia was diagnosed according to standard criteria, and details have been described previously (32-34). Supplementary Methods describe dementia surveillance and diagnosis in detail.

### 2.6 Covariates

We selected covariates a priori based on their biological relevance to gait performance or outcomes. All primary models included age (years) at baseline and sex (female or male). Models of neuropsychological performance and depressive symptoms also included education (no high school degree, high school degree, some college, or college degree). Dementia models included *apolipoprotein* ε4 (*APOE* ε4) status (carriers ≥1 ε4 allele vs non-carriers 0 ε4 alleles), derived from polymerase chain reaction-based genotyping (35, 36). Sensitivity analyses used current diabetes and the log-transformed Framingham Stroke Risk Profile (FSRP), which includes antihypertensive treatment (27).

### 2.7 Statistical Analyses

We summarized baseline characteristics as means (SD), medians (IQR), or counts (%). We evaluated cross-sectional associations between walking speed and neuropsychological test performance using multivariable linear regression, with test scores entered as continuous outcomes, and cross-sectional associations with high depressive symptoms (CES-D ≥ 16) using logistic regression. We examined associations with incident stroke and incident dementia using Cox proportional hazards regression, excluding those with prevalent events at baseline and calculating follow-up from baseline to the first occurrence of the outcome and censoring participants without the event at death, last date known free of the event, or end of 10-year follow-up, whichever came first. We conducted all analyses in the overall sample and separately in women and men. We did not test formal sex-by-exposure interactions; we report results in women and men as within-group estimates and interpret them as hypothesis-generating.

In primary analyses, we examined gait adaptability and its components, QWS and UWS, as continuous variables in separate models and expressed results per one SD decrease.

We conducted additional analyses to test the robustness and clinical interpretability of our primary findings and report the results in the Supplement. To facilitate clinical translation, we examined UWS and QWS categorically using UWS ≤0.8 m/s, a consensus threshold for poor physical performance (37), and QWS ≤1.0 m/s, the fifth-percentile cutoff used in prior work (12). We additionally adjusted all models for the FSRP and adjusted dementia models for education and current diabetes to test whether vascular risk accounted for the associations; we repeated the primary dementia models among participants with complete covariate data. Finally, we repeated analyses of cognitive performance, depressive symptoms, and incident stroke among participants aged ≥60 years at the neuropsychological examination. We report beta coefficients with standard errors for linear regression, odds ratios with 95% confidence intervals (CI) for logistic regression, and hazard ratios with 95% CI for Cox models. All statistical tests were two-sided, with significance set at p<.05. We performed analyses using SAS, version 9.4 (SAS Institute Inc).

## 3 RESULTS

### 3.1 Sample Characteristics

The larger sample comprised 2,308 participants (mean age 61.5±9.2 years; 54% women), and the dementia sample comprised 1,148 participants (mean age 68.6±5.8 years; 53% women). Tables 1a and 1b present baseline characteristics, and Table 1c presents outcome distributions. In both samples, men more often had cardiovascular disease, diabetes, and atrial fibrillation, and women more often had high depressive symptoms (11% vs. 5%).

**Table 1a.** Baseline Characteristics for Cross-sectional and Stroke Analytic Sample.

|  |  |  |  |
| --- | --- | --- | --- |
| Demographics | Total<br>(n=2308) | Women<br>(n=1247) | Men<br>(n=1061) |
| **Clinical Variables** |  |  |  |
| Age*, years, mean ± SD (range) | 61.49±9.15<br>(34.04, 88.23) | 61.22±9.15<br>(34.04, 86.23) | 61.82±9.14<br>(37.30, 88.23) |
| Female, n/total (%) | 1247/2308 (54.03%) |  |  |
| Education, n/total (%) |  |  |  |
| No high school degree | 79/2308 (3.42%) | 27/1247 (2.17%) | 52/1061 (4.90%) |
| High school degree | 630/2308 (27.30%) | 381/1247 (30.55%) | 249/1061 (23.47%) |
| Some college | 704/2308 (30.50%) | 439/1247 (35.20%) | 265/1061 (24.98%) |
| College degree | 895/2308 (38.78%) | 400/1247 (32.08%) | 495/1061 (46.65%) |
| APOE ε4 carriers, n/total (%) | 504/2235 (22.55%) | 276/1209 (22.83%) | 228/1026 (22.22%) |
| Framingham Stroke Risk Profile, median [IQR] | 1.94 [0.83, 5.18] | 1.23 [0.65, 4.00] | 2.81 [1.22, 6.98] |
| Diabetes Mellitus, n/total (%) | 224/2263 (9.90%) | 86/1221 (7.04%) | 138/1042 (13.24%) |
| Systolic blood pressure, mmHg, mean ± SD | 125.89±18.41 | 124.95±19.54 | 126.99±16.91 |
| Smoking, n/total (%) | 295/2308 (12.78%) | 166/1247 (13.31%) | 129/1061 (12.16%) |
| Atrial fibrillation, n/total (%) | 79/2308 (3.42%) | 20/1247 (1.60%) | 59/1061 (5.56%) |
| Prevalent cardiovascular disease, n/total (%) | 233/2308 (10.10%) | 83/1247 (6.66%) | 150/1061 (14.14%) |
| Coronary Heart Disease, n/total (%) | 176/2308 (7.63%) | 50/1247 (4.01%) | 126/1061 (11.88%) |
| **Walking Speed** |  |  |  |
| Quick Walking Speed, m/s, median [IQR] | 2.00 [1.33, 2.00] | 2.00 [1.33, 2.00] | 2.00 [1.33, 2.00] |
| Usual Walking Speed, m/s, median [IQR] | 1.33 [1.00, 1.33] | 1.33 [1.00, 1.33] | 1.33 [1.00, 1.33] |
| Gait Adaptability, m/s, median [IQR] | 0.67 [0.33, 0.67] | 0.33 [0.20, 0.67] | 0.67 [0.33, 0.67] |
| Quick Walking Speed ≤ 1 m/s, n/total (%) | 118/2308 (5.11%) | 73/1247 (5.85%) | 45/1061 (4.24%) |
| Usual Walking Speed ≤ 0.8 m/s, n/total (%) | 134/2308 (5.81%) | 74/1247 (5.93%) | 60/1061 (5.66%) |
| *Age is defined as age at the NP test at Exam 7, when most walking data were collected.<br>Note: All values represent mean (standard deviation) unless otherwise indicated by n (%) or median [IQR]. Walking speed was measured to the nearest second over a 4-meter course. Median and IQR values reflect the discrete distribution of possible timing values. Gait adaptability is calculated as quick walking speed minus usual walking speed.<br>Abbreviations: APOE, apolipoprotein E; NP, neuropsychological assessment |  |  |  |

**Table 1b.** Baseline Characteristics for Dementia Analytic Sample.

| **Table 1b. Baseline Characteristics for Dementia Analytic Sample** |  |  |  |
| --- | --- | --- | --- |
| Demographics | Total<br>(n=1148) | Women<br>(n=607) | Men<br>(n=541) |
| **Clinical Variables** |  |  |  |
| Age*, years, mean ± SD (range) | 68.58±5.79<br>(60.02, 88.23) | 68.51±8.83<br>(60.02, 86.23) | 68.67±5.74<br>(60.03, 88.23) |
| Female, n/total (%) | 607/1148 (52.87%) |  |  |
| Education, n/total (%) |  |  |  |
| No high school degree | 60/1148 (5.23%) | 21/607 (3.46%) | 39/541 (7.21%) |
| High school degree | 355/1148 (30.92%) | 220/607 (36.24%) | 135/541 (24.95%) |
| Some college | 348/1148 (30.31%) | 215/607 (35.42%) | 133/541 (24.58%) |
| College degree | 385/1148 (33.54%) | 151/607 (24.88%) | 234/541 (43.25%) |
| APOE ε4 carriers, n/total (%) | 259/1148 (22.56%) | 135/607 (22.24%) | 124/541 (22.92%) |
| Framingham Stroke Risk Profile, median [IQR] | 4.43 [2.30, 8.26] | 3.78 [1.69, 6.37] | 5.57 [3.10, 10.71] |
| Diabetes Mellitus, n/total (%) | 136/1126 (12.08%) | 53/598 (8.86%) | 83/528 (15.72%) |
| Systolic blood pressure, mmHg, mean ± SD | 131.04±18.73 | 130.86±19.76 | 131.25±17.51 |
| Smoking, n/total (%) | 95/1148 (8.28%) | 57/607 (9.39%) | 38/541 (7.02%) |
| Atrial fibrillation, n/total (%) | 58/1148 (5.05%) | 16/607 (2.64%) | 42/541 (7.76%) |
| Prevalent cardiovascular disease, n/total (%) | 184/1148 (16.03%) | 60/607 (9.88%) | 124/541 (22.92%) |
| Coronary Heart Disease, n/total (%) | 130/1148 (11.32%) | 36/607 (5.93%) | 94/541 (17.38%) |
| **Walking Speed** |  |  |  |
| Quick Walking Speed, m/s, median [IQR] | 1.33 [1.33, 2.00] | 1.33 [1.33, 2.00] | 2.00 [1.33, 2.00] |
| Usual Walking Speed, m/s, median [IQR] | 1.33 [1.00, 1.33] | 1.33 [1.00, 1.33] | 1.33 [1.00, 1.33] |
| Gait Adaptability, m/s, median [IQR] | 0.33 [0.33, 0.67] | 0.33 [0.33, 0.67] | 0.67 [0.33, 0.67] |
| Quick Walking Speed ≤ 1 m/s, n/total (%) | 83/1148 (7.23%) | 51/607 (8.40%) | 32/541 (5.91%) |
| Usual Walking Speed ≤ 0.8 m/s, n/total (%) | 102/1148 (8.89%) | 57/607 (9.39%) | 45/541 (8.32%) |
| **Dementia Outcome** |  |  |  |
| Incident Dementia (10-year FU), n/total (%) | 83/1148 (7.23%) | 52/607 (8.57%) | 31/541 (5.73%) |
| *Age is defined as age at the NP test at Exam 7, when most walking data were collected.<br>Note: All values represent mean (standard deviation) unless otherwise indicated by n (%) or median [IQR]. Walking speed was measured to the nearest second over a 4-meter course. Median and IQR values reflect the discrete distribution of possible timing values. Gait adaptability is calculated as quick walking speed minus usual walking speed.<br>Abbreviations: APOE, apolipoprotein E; NP, neuropsychological assessment. |  |  |  |

**Table 1c.**
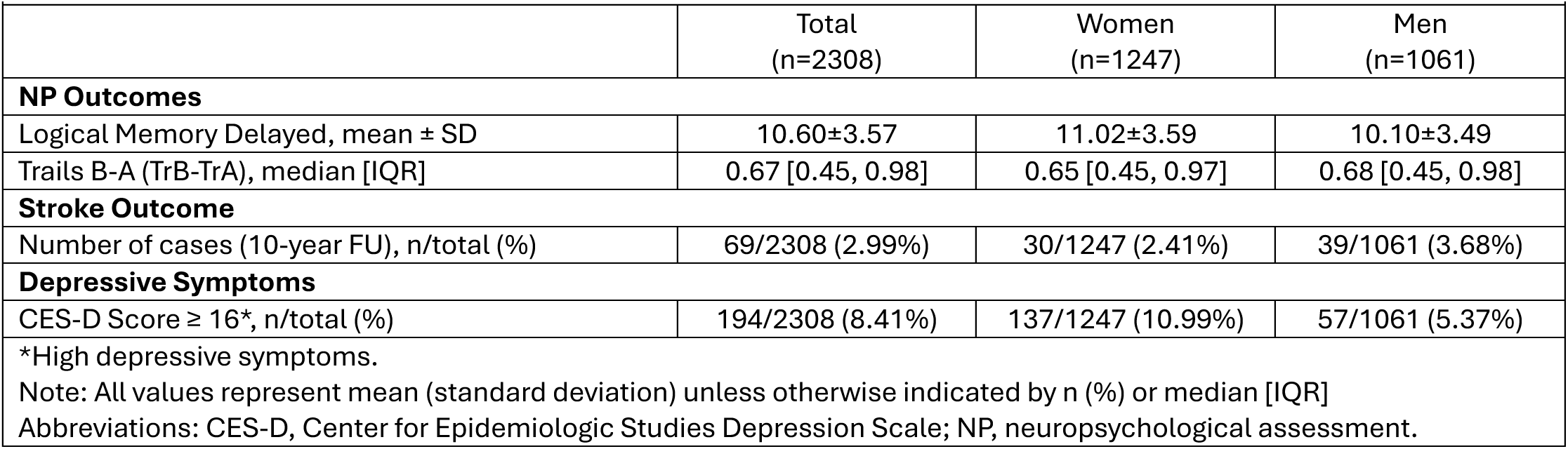
Distribution of Outcomes for Cross-sectional and Stroke Analytic Sample.

We organized results by outcome in the sections below. Associations with cognitive performance, depressive symptoms, and incident stroke are reported in Table 2, and associations with incident dementia are reported in Table 3. Categorical slow-gait analyses and sensitivity analyses adjusted for FSRP, education, and as well as restricted to participants aged ≥60 years, are reported in Tables S1 through S3.

**Table 2.** Associations of Quick Walking Speed, Usual Walking Speed, and Gait Adaptability with Neuropsychological Outcomes, Depressive Symptoms, and Incident Stroke.

|  |  |  | Quick Walk Speed<sup>2</sup><br>↓ 1 SD |  | Usual Walk Speed<sup>2</sup><br>↓ 1 SD |  | Gait Adaptability<sup>2</sup><br>↓ 1 SD |  |
| --- | --- | --- | --- | --- | --- | --- | --- | --- |
|  | Model | Cases/N | β±SE | p-value | β±SE | p-value | β±SE | p-value |
| Neuropsychological Outcomes<sup>3</sup> |  |  |  |  |  |  |  |  |
| Logical Memory Delayed | Overall | 2308 | 0.003±0.074 | 0.964 | 0.003±0.072 | 0.971 | -0.023±0.071 | 0.742 |
|  | Women | 1247 | 0.097±0.101 | 0.335 | 0.127±0.098 | 0.196 | -0.012±0.095 | 0.897 |
|  | Men | 1061 | -0.101±0.109 | 0.354 | -0.146±0.107 | 0.176 | -0.031±0.106 | 0.767 |
| Log(Trails B-A) | Overall | 2308 | **-0.020±0.004** | **<0.001** | **-0.014±0.004** | **<0.001** | **-0.012±0.004** | **0.003** |
|  | Women | 1247 | -0.011±0.006 | 0.053 | -0.002±0.005 | 0.753 | **-0.012±0.005** | **0.029** |
|  | Men | 1061 | **-0.031±0.006** | **<0.001** | **-0.028±0.006** | **<0.001** | **-0.014±0.006** | **0.030** |
|  |  |  | OR (95% CI) | p-value | OR (95% CI) | p-value | OR (95% CI) | p-value |
| High Depressive Symptoms<br>(CES-D ≥ 16)<sup>3</sup> | Overall | 2308 | **1.24 (1.07, 1.45)** | **0.005** | **1.26 (1.09, 1.46)** | **0.002** | 1.04 (0.90, 1.20) | 0.627 |
|  | Women | 1247 | **1.23 (1.02, 1.47)** | **0.027** | **1.24 (1.04, 1.48)** | **0.016** | 1.03 (0.87, 1.23) | 0.740 |
|  | Men | 1061 | 1.26 (0.95, 1.67) | 0.111 | 1.29 (0.98, 1.69) | 0.075 | 1.06 (0.80, 1.40) | 0.700 |
|  |  |  | HR (95% CI) | p-value | HR (95% CI) | p-value | HR (95% CI) | p-value |
| Incident Stroke, 10-year-follow-up | Overall | 69/2308 | **1.30 (1.03, 1.64)** | **0.025** | **1.36 (1.11, 1.65)** | **0.003** | 0.96 (0.75, 1.24) | 0.772 |
|  | Women | 30/1247 | 1.09 (0.76, 1.56) | 0.627 | 1.13 (0.80, 1.59) | 0.484 | 0.96 (0.66, 1.41) | 0.849 |
|  | Men | 39/1061 | **1.55 (1.11, 2.17)** | **0.011** | **1.68 (1.23, 2.29)** | **0.001** | 0.96 (0.68, 1.35) | 0.814 |

Note: Gait adaptability is calculated as quick walking speed minus usual walking speed.
<sup>1</sup> Overall model adjusted for age and sex; sex-specific model adjusted for age only.
<sup>2</sup>Usual walking speed, Quick walking speed, and gait adaptability were natural log-transformed to normalize their distributions, standardized, and multiplied by -1 so higher values reflect slower walking or a smaller QWS-UWS margin.
<sup>3</sup>Adjusted for education in addition to age and sex.

**Table 3.** Associations of Quick Walking Speed, Usual Walking Speed, and Gait Adaptability with Incident Dementia.

| Outcome |  |  | Quick Walk Speed<sup>2</sup><br>↓ 1 SD |  | Usual Walk Speed<sup>2</sup><br>↓ 1 SD |  | Gait Adaptability<sup>2</sup><br>↓ 1 SD |  |
| --- | --- | --- | --- | --- | --- | --- | --- | --- |
|  | Model | Cases/N | HR (95% CI) | p-value | HR (95% CI) | p-value | HR (95% CI) | p-value |
| Incident Dementia, 10 Year Follow Up | Overall | 83/1148 | 1.34 (1.10, 1.63) | 0.004 | 1.20 (0.98, 1.46) | 0.078 | 1.20 (0.98, 1.47) | 0.085 |
|  | Women | 52/607 | 1.33 (1.06, 1.66) | 0.014 | 1.14 (0.88, 1.47) | 0.314 | 1.35 (1.04, 1.76) | 0.023 |
|  | Men | 31/541 | 1.37 (0.91, 2.05) | 0.132 | 1.36 (0.92, 1.99) | 0.121 | 0.96 (0.66, 1.41) | 0.835 |

Note: Gait adaptability is calculated as quick walking speed minus usual walking speed.
<sup>1</sup> Overall model adjusted for age, sex, and APOE ε4 status; sex-specific models adjusted for age and APOE ε4 status.
<sup>2</sup>Quick walking speed, usual walking speed, and gait adaptability were natural log-transformed to normalize their distributions, standardized, and multiplied by -1 so higher values reflect slower walking or a smaller QWS-UWS margin.

### 3.2 Gait Speed and Cognitive Performance

Both slower QWS and UWS were associated with worse performance on Trails B-A (QWS: β±SE=−0.020±0.004, p<0.001; UWS: −0.014±0.004, p<0.001). These estimates correspond to roughly 1% to 2% longer Trails B-A completion time per SD slower walking. Reduced gait adaptability was also associated with worse Trails B-A performance (−0.012±0.004, p=0.003). Neither QWS, UWS, nor gait adaptability was associated with Logical Memory Delayed Recall.

### 3.3 Gait Speed and Depressive Symptoms

Slower QWS and UWS were associated with higher odds of high depressive symptoms (QWS: OR 1.24, 95% CI 1.07–1.45, p=0.005; UWS: OR 1.26, 95% CI 1.09–1.46, p=0.002). Gait adaptability was not associated with depressive symptoms.

### 3.4 Gait Speed and Incident Stroke

Over 10 years, 69 participants (3.0%) had an incident stroke. Each one-standard deviation decrease in QWS was associated with a higher hazard of incident stroke (HR 1.30, 95% CI 1.03–1.64, p=0.025), with a similar association observed for UWS (HR 1.36, 95% CI 1.11–1.65, p=0.003). Gait adaptability was not associated with incident stroke.

### 3.5 Gait Speed and Incident Dementia

Over 10 years, 83 of the 1,148 participants with complete *APOE* ε4 genotyping developed dementia. Each one-standard deviation decrease in QWS was associated with a higher hazard of incident dementia (HR 1.34, 95% CI 1.10–1.63, p=0.004), whereas UWS was not. Gait adaptability was not associated with incident dementia.

### 3.6 Associations in Women and Men

#### 3.6.1 Cognitive Performance

Slower QWS and UWS were associated with worse Trails B-A performance in men (QWS: −0.031±0.006, p<0.001; UWS: −0.028±0.006, p<0.001) but not in women. Gait adaptability was associated with worse Trails B-A performance in both women (−0.012±0.005, p=0.029) and men (−0.014±0.006, p=0.030). Neither walking speed nor gait adaptability was associated with Logical Memory Delayed Recall in women or men.

#### 3.6.2 Depressive Symptoms

Slower QWS and UWS were associated with higher odds of high depressive symptoms in women (QWS: OR 1.23, 95% CI 1.02–1.47, p=0.027; UWS: OR 1.24, 95% CI 1.04–1.48, p=0.016) but not in men, although estimates in men were similar in size (QWS OR 1.26; UWS OR 1.29) and based on fewer cases (57 vs 137). Gait adaptability was not associated with depressive symptoms in either women or men.

#### 3.6.3 Incident Stroke

Slower QWS and UWS were associated with higher hazard of incident stroke in men (QWS: HR 1.55, 95% CI 1.11–2.17, p=0.011; UWS: HR 1.68, 95% CI 1.23–2.29, p=0.001) but not in women. Gait adaptability was not associated with incident stroke in either women or men.

#### 3.6.4 Incident Dementia

In women (n=607, 52 incident cases), slower QWS was associated with incident dementia (HR 1.33, 95% CI 1.06–1.66, p=0.014), whereas UWS was not. Gait adaptability was associated with incident dementia in women (HR 1.35, 95% CI 1.04–1.76, p=0.023). In men (n=541, 31 incident cases), neither QWS nor UWS was associated with incident dementia, although both estimates were similar to the QWS estimate in women. Gait adaptability was not associated with incident dementia in men.

### 3.7 Sensitivity Analyses

Executive function and depressive symptom findings were generally similar after FSRP adjustment and age restriction (≥60 years). However, the gait adaptability–executive function association in men no longer reached significance in either analysis, nor did the QWS–depressive symptom association in women after FSRP adjustment (Tables S1, S3).

After FSRP adjustment, stroke associations with QWS and UWS remained significant in men; after age restriction (≥60 years), they remained significant overall and in men (Tables S1 and S3). Dementia associations with QWS overall and in women, and with gait adaptability in women, persisted after additional adjustment (Table S2).

## 4 DISCUSSION

In this prospective community-based study, slower QWS and UWS were similarly associated with poorer executive function, higher odds of high depressive symptoms, and greater risk of incident stroke. The positive association between QWS and incident dementia was statistically significant, and the UWS association was not, although the two confidence intervals overlapped. Gait adaptability, the difference between the two paces, was associated with executive function in women and men and with incident dementia in women, but not with depressive symptoms, stroke, or verbal memory. Together, these findings show that a study recording usual pace alone would not have observed the dementia and adaptability associations reported here, with implications for how gait is assessed.

### 4.1 Quick versus Usual Walking Speed for Incident Dementia

Slower QWS was associated with higher 10-year dementia risk, consistent with earlier work in this cohort linking quick-pace walking to incident dementia (12). The UWS estimate was smaller and not statistically significant. In the UK Biobank, maximal walking speed from wrist-worn sensors likewise showed a stronger association with dementia than UWS (13), suggesting that the pattern extends beyond clinic-based stopwatch timing. Quick walking may capture dementia-relevant neuromotor vulnerability that habitual pace masks because accelerating above self-selected pace engages volitional motor control and cognitive-motor integration through frontal-subcortical circuitry (4), whereas habitual walking is largely automated in healthy adults (38).

Consistent with this interpretation, neuroimaging evidence shows that walking fast recruits greater prefrontal and motor cortex activation in older adults than in younger adults (39). In a memory clinic sample (the GAIT study), worse cognitive stage was associated with slower fast and preferred walking speeds and a smaller walking speed reserve, a measure that parallels our adaptability construct (40).

### 4.2 Associations in Women and Men

We repeated primary analyses separately in women and men and did not test formal sex-by-exposure interactions. The estimates therefore describe within-group patterns and are hypothesis-generating.

For incident dementia over up to 10-years of follow-up, slower QWS and lower gait adaptability were associated with higher risk in women, while UWS was not; in men, no gait measure reached significance. The QWS estimates were nearly identical in women and men (HR 1.33 and 1.37), so the difference in significance reflects fewer cases in men (31 vs 52). The adaptability estimates differed in direction (HR 1.35 in women, 0.96 in men), though their confidence intervals overlapped. For adaptability, one untested possibility is that women maintain habitual pace as neural capacity declines but lose the ability to accelerate on demand, consistent with a cognitive reserve interpretation (41). For incident stroke and executive function, QWS and UWS associations were significant in men but not in women, a pattern consistent with earlier accumulation of cardiovascular and cerebrovascular burden in men (16), although only 30 strokes occurred in women. By contrast, the association between lower adaptability and worse executive function was similar in women and men.

### 4.3 Gait as a Transdiagnostic Marker of Brain Health

For executive function, depressive symptoms, and incident stroke, QWS and UWS were similarly informative, consistent with slower gait indexing shared frontal-subcortical and vascular substrates that give rise to multiple brain conditions (42), rather than a mechanism specific to any one outcome. Gait adaptability showed a narrower pattern, with associations with executive function and incident dementia in women over 10 years, but not with incident stroke or depressive symptoms. The adaptability intervals for stroke (HR 0.96, 95% CI 0.75-1.24) and depressive symptoms (OR 1.04, 95% CI 0.90-1.20) excluded the effect sizes observed for either pace, indicating precise null findings rather than underpowered estimates. Adjustment for vascular risk attenuated the stroke associations more than the executive function and depressive symptom associations, suggesting that vascular burden accounts for much of the link between walking speed and stroke. The co-occurrence of gait slowing, executive dysfunction, and depressive symptoms in older adults has been described as a distinct clinical phenotype linked to frontal-subcortical circuitry and vascular pathology (9), consistent with our findings.

Neither walking speed nor gait adaptability was associated with delayed recall, consistent with prior research linking gait to frontal-subcortical rather than medial temporal function (4, 42), and with longitudinal evidence that the pace domain of gait predicts decline in executive function (6). An earlier analysis of this cohort, restricted to participants with brain MRI and covering a broader cognitive battery, linked slower gait to poorer verbal memory (12). Our confidence intervals excluded differences larger than 0.17 points, a precise null rather than an underpowered one. This null does not conflict with the dementia findings. Cross-sectional test performance and 10-year risk of a clinical diagnosis are different constructs, and the executive function associations we observed offer a route to dementia that does not run through episodic memory. One interpretive distinction is, however, the dementia outcome. Earlier Framingham work linked quick walking with clinically diagnosed Alzheimer’s disease (12), whereas we examined incident dementia overall. With 83 cases we could not assess whether the QWS association varies by etiology, which future studies should address..

### 4.4 Research and Clinical Implications

Adding a quick walking trial requires little time and no additional equipment, enabling assessment of quick pace and adaptability. Whether adaptability identifies dementia risk among people with normal UWS warrants direct testing.

Detection timing may matter as much as cost. In the dementia sample (mean age 68.5 years), 91% walked faster than the 0.8 m/s slow-gait threshold at usual pace, so a reduced capacity to accelerate may flag cognitive risk while usual pace still looks unremarkable. Analyses restricted to people with normal usual pace could test this directly.

Recording both paces with finer resolution and the same number of trials would sharpen the measure at almost no added cost, with wearable sensors extending this assessment beyond the clinic (13). Further work will require joint modeling of QWS and UWS, formal tests of sex interactions, replication in diverse populations, and inclusion of neuroimaging or fluid biomarkers to identify underlying pathways.

### 4.5 Strengths and Limitations

Evaluating both paces within the same cohort allowed a direct comparison unavailable to studies recording usual pace alone. Examining five outcomes in one cohort allowed cross-outcome comparisons, and rigorous surveillance supported high-confidence ascertainment of incident dementia and stroke.

However, we did not formally evaluate whether QWS or gait adaptability improved dementia prediction beyond UWS alone, and we did not test sex-by-exposure interactions. Furthermore, whole-second timing and unequal numbers of trials likely attenuated the estimates, particularly for gait adaptability, an absolute difference that partly scales with walking speed. We assessed gait only once, and cross-sectional cognitive and depressive symptom analyses preclude causal inference. Finally, we did not model death as a competing risk, and the predominantly white European ancestry of the sample limits generalizability.

### 4.6 Conclusions

Both UWS and QWS were associated with executive function, depressive symptoms, and incident stroke. QWS was also associated with incident dementia over 10 years, whereas the UWS association did not reach statistical significance. Gait adaptability was associated with executive function and with incident dementia in women over 10 years. These findings support further evaluation of a brief quick-walk trial alongside usual-pace assessment to capture gait characteristics relevant to brain health.

## Data Availability

All data produced in the present study are available upon reasonable request to the authors.

https://www.framinghamheartstudy.org

## ACKNOWLEDGMENTS

We thank the research team at Boston University and all Framingham Heart Study participants who volunteered their time and data and made this scientific effort possible.

## CONSENT STATEMENT

All study participants signed the written informed consent form.

