## Supplementary for "Beyond Usual Pace: Quick Walking Speed and Brain Health Outcomes in the Framingham Heart Study"

### Methods

#### Dementia Surveillance and Diagnosis

Cognitive status was monitored at each clinical examination using the Mini-Mental State Examination (MMSE) (22-24). Participants were identified for further evaluation through a multi-source surveillance approach that incorporated education-adjusted MMSE performance, prior FHS examination data, self and family-reported cognitive concerns, primary care and specialist referrals, hospital and stroke surveillance record, and regular neuropsychological testing (22-24). Those identified were referred for further evaluation.

Participants with suspected cognitive impairments were followed longitudinally by a neurologist and neuropsychologist, while those with suspected dementia were reviewed by a clinical multidisciplinary panel. Diagnosis, subtype and estimated diagnosis date were established through serial clinical evaluations, caregiver interviews, medical records, neuroimaging, and autopsy findings were available. Medical records were also reviewed postmortem for cognitive decline occurring after the final clinical examination, and cases diagnosed before 2001 were re-reviewed against current diagnostic criteria.

Dementia was diagnosed according to standard criteria as described previously (25-27). Including established criteria for Alzheimer disease, vascular dementia and dementia with Lewy bodies.

**Supplementary Table S1. Associations of Quick Walking Speed, Usual Walking Speed, and Gait Adaptability with Neuropsychological Outcomes, Depressive Symptoms, and Incident Stroke adjusted by Framingham Stroke Risk Profile**

| | | | Quick Walk Speed[2] ↓ 1 SD | | Usual Walk Speed[2] ↓ 1 SD | | Gait Adaptability[2] ↓ 1 SD | |
|---|---|---|---|---|---|---|---|---|
| | **Model** | **Cases/N** | **β±SE** | **p-value** | **β±SE** | **p-value** | **β±SE** | **p-value** |
| **Neuropsychological Outcomes[3]** | | | | | | | | |
| Logical Memory Delayed | Overall | | 0.000±0.076 | 1.000 | -0.013±0.074 | 0.856 | -0.012±0.071 | 0.863 |
| | Women | | 0.102±0.104 | 0.323 | 0.115±0.100 | 0.250 | 0.004±0.096 | 0.965 |
| | Men | | -0.111±0.111 | 0.314 | -0.161±0.109 | 0.138 | -0.028±0.107 | 0.795 |
| Log(Trails B-A) | Overall | | **-0.019±0.004** | **<0.001** | **-0.013±0.004** | **0.002** | **-0.011±0.004** | **0.005** |
| | Women | | -0.011±0.006 | 0.062 | -0.001±0.006 | 0.884 | **-0.012±0.005** | **0.027** |
| | Men | | **-0.028±0.006** | **<0.001** | **-0.027±0.006** | **<0.001** | -0.011±0.006 | 0.065 |
| | | | **OR (95% CI)** | **p-value** | **OR (95% CI)** | **p-value** | **OR (95% CI)** | **p-value** |
| **High Depressive Symptoms (CES-D≥16)[3]** | Overall | | **1.19 (1.02, 1.40)** | **0.030** | **1.22 (1.05, 1.43)** | **0.011** | 1.02 (0.88, 1.19) | 0.784 |
| | Women | | 1.21 (0.99, 1.46) | 0.058 | **1.21 (1.01, 1.46)** | **0.042** | 1.03 (0.86, 1.23) | 0.750 |
| | Men | | 1.16 (0.86, 1.55) | 0.333 | 1.23 (0.92, 1.63) | 0.164 | 1.00 (0.74, 1.35) | 1.000 |
| | | | **HR (95% CI)** | **p-value** | **HR (95% CI)** | **p-value** | **HR (95% CI)** | **p-value** |
| **Incident Stroke, 10 Year Follow Up** | Overall | 66/2262 | 1.14 (0.89, 1.47) | 0.300 | 1.23 (0.97, 1.56) | 0.081 | 0.90 (0.69, 1.18) | 0.453 |
| | Women | 29/1220 | 0.88 (0.60, 1.29) | 0.507 | 0.93 (0.64, 1.33) | 0.688 | 0.88 (0.59, 1.32) | 0.543 |
| | Men | 37/1042 | **1.42 (1.00, 2.00)** | **0.049** | **1.52 (1.11, 2.07)** | **0.009** | 0.92 (0.64, 1.33) | 0.657 |

Note: Gait adaptability is calculated as quick walking speed minus usual walking speed.

[1] Overall Model: adjusted for sex; sex-specific model: did not adjust for sex. FSRP was log transformed to normalize distribution.

[2]Usual Walk Speed, Quick Walk Speed, and Gait Adaptability are natural log transformed to normalize their distributions, standardized, and then multiplied by -1.

[3]Adjusted for education.

**Supplementary TableS2. Associations between Walk Speed and Dementia, adjusted for multiple additional covariates**

| Adjustment | Outcome | Model | Cases/N | Quick Walk Speed[2] ↓ 1 SD HR (95% CI) | Quick Walk Speed[2] ↓ 1 SD p-value | Usual Walk Speed[2] ↓ 1 SD HR (95% CI) | Usual Walk Speed[2] ↓ 1 SD p-value | Gait Adaptability[2] ↓ 1 SD HR (95% CI) | Gait Adaptability[2] ↓ 1 SD p-value |
|---|---|---|---|---|---|---|---|---|---|
| **Primary Model+ FSRP (Full Sample)** | **Incident Dementia, 10 Year Follow Up** | Overall | **81/1126** | **1.30 (1.05, 1.60)** | **0.015** | 1.16 (0.94, 1.43) | 0.178 | 1.18 (0.95, 1.46) | 0.130 |
| | | Women | **52/598** | **1.29 (1.01, 1.64)** | **0.045** | 1.08 (0.82, 1.43) | 0.565 | **1.34 (1.03, 1.74)** | **0.029** |
| | | Men | **29/528** | 1.33 (0.86, 2.04) | 0.199 | 1.30 (0.89, 1.90) | 0.179 | 0.92 (0.62, 1.38) | 0.694 |
| **Primary Model+ Education+ Current Diabetes (Full Sample)** | **Incident Dementia, 10 Year Follow Up** | Overall | **81/1126** | **1.32 (1.08, 1.62)** | **0.006** | 1.16 (0.95, 1.43) | 0.155 | 1.20 (0.97, 1.47) | 0.089 |
| | | Women | **52/598** | **1.31 (1.04, 1.65)** | **0.021** | 1.14 (0.88, 1.47) | 0.335 | **1.33 (1.02, 1.73)** | **0.038** |
| | | Men | **29/528** | 1.35 (0.88, 2.09) | 0.172 | 1.31 (0.88, 1.96) | 0.187 | 0.96 (0.65, 1.44) | 0.856 |
| **Primary Model (Sample Restricted to Participants with Current Diabetes/FSRP Data)** | **Incident Dementia, 10 Year Follow Up** | Overall | **81/1126** | **1.35 (1.11, 1.64)** | **0.003** | 1.19 (0.97, 1.46) | 0.097 | 1.22 (0.99, 1.49) | 0.064 |
| | | Women | **52/598** | **1.34 (1.07, 1.68)** | **0.012** | 1.14 (0.89, 1.48) | 0.306 | **1.37 (1.05, 1.77)** | **0.019** |
| | | Men | **29/528** | 1,38 (0.90, 2.12) | 0.140 | 1.33 (0.90, 1.96) | 0.149 | 0.96 (0.65, 1.43) | 0.850 |

Note: Gait adaptability is calculated as quick walking speed minus usual walking speed.

[1] Overall Model: adjusted for sex; sex-specific model: did not adjust for sex. FSRP was log transformed to normalize the distribution.

[2]Usual Walk Speed, Quick Walk Speed, and Difference between Quick Walk and Usual Walk are natural log transformed to normalize their distributions, standardized, and then multiplied by -1.

| **Supplementary Table S3. Associations between Slower Walking Speed (Quick Walk and Usual Walk) and Various Outcomes, among Participants At Least 60 Years Old** | | | | | | | | |
|---|---|---|---|---|---|---|---|---|
| | | | Quick Walk Speed[2] ↓ 1 SD | | Usual Walk Speed[2] ↓ 1 SD | | Gait Adaptability[2] ↓ 1 SD | |
| | **Model** | **Cases/N** | **β±SE** | **p-value** | **β±SE** | **p-value** | **β±SE** | **p-value** |
| **Neuropsychological Outcomes[3]** | | | | | | | | |
| Logical Memory Delayed | Overall | | 0.000±0.102 | 0.998 | 0.061±0.098 | 0.539 | -0.114±0.103 | 0.269 |
| | Women | | 0.216±0.146 | 0.141 | 0.260±0.140 | 0.064 | -0.023±0.155 | 0.883 |
| | Men | | -0.218±0.143 | 0.126 | -0.153±0.139 | 0.271 | -0.186±0.137 | 0.175 |
| Log(Trails B-A) | Overall | | **-0.026±0.0006** | **<0.001** | **-0.020±0.006** | **0.001** | **-0.016±0.007** | **0.015** |
| | Women | | -0.017±0.009 | 0.052 | -0.005±0.008 | 0.561 | **-0.188±0.009** | **0.045** |
| | Men | | **-0.036±0.009** | **<0.001** | **-0.036±0.009** | **<0.001** | -0.013±0.009 | 0.143 |
| | | | **OR (95% CI)** | **p-value** | **OR (95% CI)** | **p-value** | **OR (95% CI)** | **p-value** |
| **High Depressive Symptoms (CES-D≥16)[3]** | Overall | | **1.29 (1.05, 1.51)** | **0.015** | **1.26 (1.04, 1.53)** | **0.022** | 1.14 (0.92, 1.41) | 0.244 |
| | Women | | 1.21 (0.94, 1.56) | 0.144 | 1.19 (0.94, 1.51) | 0.149 | 1.10 (0.82, 1.47) | 0.518 |
| | Men | | 1.43 (0.97, 2.11) | 0.072 | 1.35 (0.93, 1.96) | 0.114 | 1.19 (0.86, 1.64) | 0.306 |
| | | | **HR (95% CI)** | **p-value** | **HR (95% CI)** | **p-value** | **HR (95% CI)** | **p-value** |
| **Incident Stroke, 10 Year Follow Up** | Overall | 52/1231 | **1.43 (1.12, 182)** | **0.005** | **1.40 (1.14, 1.73)** | **0.002** | 1.04 (0.78, 1.38) | 0.804 |
| | Women | 23/640 | 1.18 (0.81, 1.74) | 0.394 | 1.18 (0.82, 1.71) | 0.371 | 1.00 (0.64, 1.55) | 0.984 |
| | Men | 29/591 | **1.80 (1.22, 2.66)** | **0.003** | **1.81 (1.27, 2.57)** | **0.001** | 1.06 (0.74, 1.52) | 0.749 |

Note: Gait adaptability is calculated as quick walking speed minus usual walking speed.

[1] Overall Model: adjusted for sex; sex-specific model: did not adjust for sex. FSRP was log transformed to normalize distribution.

[2]Usual Walk Speed, Quick Walk Speed, and Difference between QW and US are natural log transformed to normalize their distributions, standardized, and then multiplied by -1.

[3]Adjusted for education
